# A multi-population-based genomic analysis uncovers unique haplotype variants and crucial mutant genes in SARS-CoV-2

**DOI:** 10.1101/2022.09.05.22279597

**Authors:** Afzal Sheikh, He Huang, Sultana Parvin, Mohammad Badruzzaman, Tofayel Ahmed, Ekhtear Hossain, Iri Sato Baran, Zahangir Alam Saud

**Affiliations:** Department of Biochemistry and Molecular Biology, Bangabandhu Sheikh Mujibur Rahman Agricultural University, Gazipur 1706, Dhaka, Bangladesh; Research and Development department, Bioengineering Lab. Co., Ltd. Address: 657 Nagatake Midori-ku,Sagamihara-shi, Kanagawa-ken 252-0154 Japan; Department of Biology, Faculty of Science and Engineering, Saitama University, Saitama, Japan; Department of Agroforestry and Environment, Bangabandhu Sheikh Mujibur Rahman Agricultural University, Gazipur 1706, Dhaka, Bangladesh; Department of Biological Sciences and chemistry, Southern University and A&M College, 244 William James Hall, Baton Rouge, LA70813, USA; Genesis Institute of Genetic Research, Genesis Healthcare Corporation, Yebisu Garden Place Tower 15F/26F 4-20-3 Ebisu, Shibuya-ku, Tokyo, Japan; Department of Biochemistry and Molecular Biology, University of Rajshahi, Rajshahi 6205, Bangladesh

**Keywords:** SARS-CoV-2, genome, variants, haplotype, nucleocapsid

## Abstract

**Background:** COVID-19 is a disease caused by severe acute respiratory syndrome coronavirus 2 (SARS-CoV-2). Rigorous detection and treatment strategies against SARS-CoV-2 have become very challenging due to continuous evolutions to the viral genome. Therefore, careful genomic analysis is sorely needed to understand transmission, the cellular mechanism of pathogenicity, and the development of vaccines or drugs.

**Objective:** In this study, we intended to identify SARS-CoV-2 genome variants that may help understand the cellular and molecular foundation of coronavirus infections required to develop effective intervention strategies.

**Methods:** SARS-CoV-2 genome sequences were downloaded from an open-source public database, processed, and analyzed for variants in target detection sites and genes.

**Results:** We have identified six unique variants, G---AAC, T---AAC---T; AAC---T; C----C; C-------C; and C--------T at the nucleocapsid region and eleven major hotspot mutant genes: nsp3, surface glycoprotein, nucleocapsid phosphoprotein, ORF8, nsp6, nsp2, nsp4, helicase, membrane glycoprotein, 3’-5’ exonuclease, and 2’-O-ribose methyltransferases. In addition, we have identified eleven major mutant genes that may have a crucial role in SARS-CoV-2 pathogenesis.

**Conclusion:** Studying haplotype variants and 11 major mutant genes to understand the mechanism of action of fatal pathogenicity and inter-individual variations in immune responses is inevitable for managing target patient groups with identified variants and developing effective anti-viral drugs and vaccines.

## Background

The new case of SARS-CoV-2 outside China was first announced by the Director-General of the WHO on February 26, 2020, and is now officially known as COVID-19 disease (1). The human COVID-19 pandemic disease caused by the infections of severe acute respiratory syndrome coronavirus-2 (SARS-CoV-2), which impacts the lower respiratory tract, has spread across the globe in diverse methods and speed (2–4). The spectrum of symptoms ranges from developing mild to moderate respiratory illness that recovers without hospitalization to the lethal form of COVID-19 associated with severe pneumonia, difficulty in breathing or shortness of breath, chest pain, loss of speech or movement, and fatality (3–7).

Structurally, SARS-CoV-2 is an enveloped, 5’-capped, single-stranded polyadenylated positive-strand RNA virus of a non-segmented genome of ∼29.7 kb long encoding 16 non-structural proteins (NSPs), which are required for virus replication and pathogenesis. Four structural proteins, including envelope (E), membrane (M), nucleocapsid (N), and spike glycoprotein (S), are essential for virus subtyping, structural rearrangement of the RNA genome, assembly, budding, viral replication, pathogenesis, response to vaccines, and viral entry to host. Moreover, nine others are accessory factors that facilitate the unwinding of dsRNA, viral RNA cap formation, exonuclease activity, membrane fusion, interaction with host cells, and immune response to the host (8–12). Thus, mutations in these genes may interfere with changing proteins structures, RNA dimerization, and alterations in the functions as mentioned earlier, including interaction with RNA and signaling events (13–15). Moreover, some functional features of these genes are yet to be discovered.

To date, many drugs have been applied to manage COVID-19 patients, along with several vaccines. Unfortunately, there are no effective drugs so far, and if some of the drugs are functioning with some adverse side effects, individual patient groups are not responding to those drugs (16–22). In addition, scientific communities are aware of some repeatedly reported limitations of the already available vaccines, including recurrence of infections after being vaccinated with multiple doses, adverse side effects, and fatality (23–27). These limitations are reported in a particular group of patients while some other target groups effectively responded to those already available drugs or/ and vaccines (16–26). Therefore, in order to develop new effective therapeutic strategies for these non-responsive patient groups and adverse drug effects, it is crucial to study the association of the target variants with pathogenicity, replication rate, recurrence infections, response to host immunity, and target drugs at the cellular and molecular level.

In the present study, we focused on characterizing the accumulation of mutations and a detailed understanding of the geographic distribution of genetic variants in 1,012,582 sequences, including 405,461 complete genome sequences from the NCBI database as of August 4, 2021. From the shreds of evidence, we are reporting for the first time the seven unique haplotype variants in the nucleocapsid region, four of which is in the target RT-PCR detection sites recommended by the central research institute CDC in the USA, China, Germany, and Japan’s Center of Infectious Disease (NIID) testing protocol (28–30). In addition, we have identified the major hotspot mutant genes, some of which have been reported before to be associated with RNA capping and viral replication, infection, and pathogenesis. Therefore, this study will be of great interest to scientists working in cellular and molecular biology, molecular pathogenicity, medicine, and researchers working in vaccine development, including the scientific community working on infectious diseases detection, diagnosis methods, and human health care.

## Methods

### SARS-CoV-2 Sequence data

We intended to analyze the major hotspot mutations at the nucleocapsid phosphoprotein and envelop region since these two regions are the major target for RT-PCR-based detection of COVID-19 positive cases by CDC in the USA, China, Germany, and NIID in Japan. In addition, major hotspot mutant sites were analyzed for the complete genome of COVID-19 sequence data of global samples from the open sources database. Therefore, we first downloaded 1,012,582 available SARS-CoV-2 global sequence data from the National Center for Biotechnology Information (NCBI) database. We then separately processed and analyzed the complete and partial sequence data.

### Data processing

After downloading the sequence, data were processed for variant analysis using Linux terminal using the following command lines, python and muscle program for target region, N and E (nucleocapsid phosphoprotein and envelope protein)

grep “>” covid_19.fasta | grep nucleocapsid > goi.txt

grep “>” covid_19.fasta | grep “nucelocapsid” | sed-e’s/>//g’ |cut -d “ ” -f1 > nucleocapsid.list

Create_sorted fasta file (nucleocapsid gene):

python3 pull_fasta.py -f covid_19.fasta -l nucleocapsid.list > nucleocapsid.fasta

Collapse duplicate sequences into single sequence:

Vsearch --derep_fulllength nucleocapsid_covid-19.fasta --output uniquenucleocapsid_covid-19.fasta

### Sequence alignment and mutation analysis

The commands were performed repeatedly for each target gene. The obtained sequences were further analyzed using muscle for the alignment and to separate unique sequences against the reference Covid-19 genome, NC_045512 from Wuhan in China. The command-line used is as follows:

muscle -in uniqenvelop_Covid-19.fasta -out uniqenvelopseq.fasta_alingnedseq.

The unique aligned sequences are then used for the analysis of variation/mutation using jalview application.

In addition, we also analyzed the hotspot mutation sites towards the complete genome of SARS-CoV-2 based on our data filtering criteria stated above and using “View Mutations” in the SARS-CoV-2 SRA Data” link.

(https://www.ncbi.nlm.nih.gov/labs/virus/vssi/#/virus?SeqType_s=Nucleotide&Viru_sLineage_ss=SARS-CoV-2,%20taxid:2697049). The data obtained were then analyzed for each gene and mutations, including the protein change type (synonymous/ non-synonymous)

## Results

### Unique SARS-CoV-2 clones were identified with mutations at the target detection sites in global samples

Since we identified false-negative results in ∼16% of the covid positive patients, which were confirmed using several primer sets, we intensely wanted to investigate variations in the SARS-CoV-2 genome, particularly the region where primer-probe sets are designed and recommended by CDC, USA; NIID, Japan, CDC China, Germany and others (28–30). We have identified many global samples which have multiple mutations at the same primer-probe binding site. Some clones were identified to have mutations at both primer and probe binding sites. Some were identified to have mutations at multiple primers-probe binding sites, while some global samples were found to have mutations at either of the two primers or probe binding sites (Table 1).

**Table 1.** The number of global samples identified with the mutation at the primer-probe binding sites for RT-PCR-based detection of severe acute respiratory syndrome coronavirus 2 (SARS-CoV-2).

| SARS-CoV-2 | PCR detection sites (primers and probes) |  |  |  |  |  |  |
| --- | --- | --- | --- | --- | --- | --- | --- |
|  | US_CDC_N2 | JP_NIID_N2 | China_CDC_N2 | JP_NIID_2<br>and<br>US_CDC_N2 | China_CDC_N2<br>and JP_NIID_N2 | China_CDC_N2<br>and US_CDC_N2 | JP_NIID_2;<br>China_CDC_N2<br>and US_CDC_N2 |
| Number of clones with mutation at the detection sites | 52 | 44 | 60 | 10 | 10 | 17 | 1 |

We speculated in the previous study that mutation at the target detection site might significantly impact false-negative results, which we reported is ∼16% in Japanese samples (30). Our present results also indicate the essence of using multiple (at least three) primer sets to reduce the transmission of SARS-CoV-2 infections rate caused by false-negative results.

### SARS-CoV-2 clones with unique haplotype variants are present in the nucleocapsid region

While analyzing the processed data, we identified six unique haplotype mutation patterns G-----AAC, T-----AAC-----T; AAC---T; C----C; C-------C; and C--------T present in the nucleocapsid region of the SARS-CoV-2 genome (Figure 1. a) – d)). No similar haplogroup pattern could be identified at other RT-PCR target detection sites. Furthermore, although not at the target detection (primer-probe binding site) sites, three different haplotype variants were also observed ((Figure 1. e) - f)) in the nucleocapsid region that encodes for nucleocapsid phosphoproteins.

**Figure 1.**
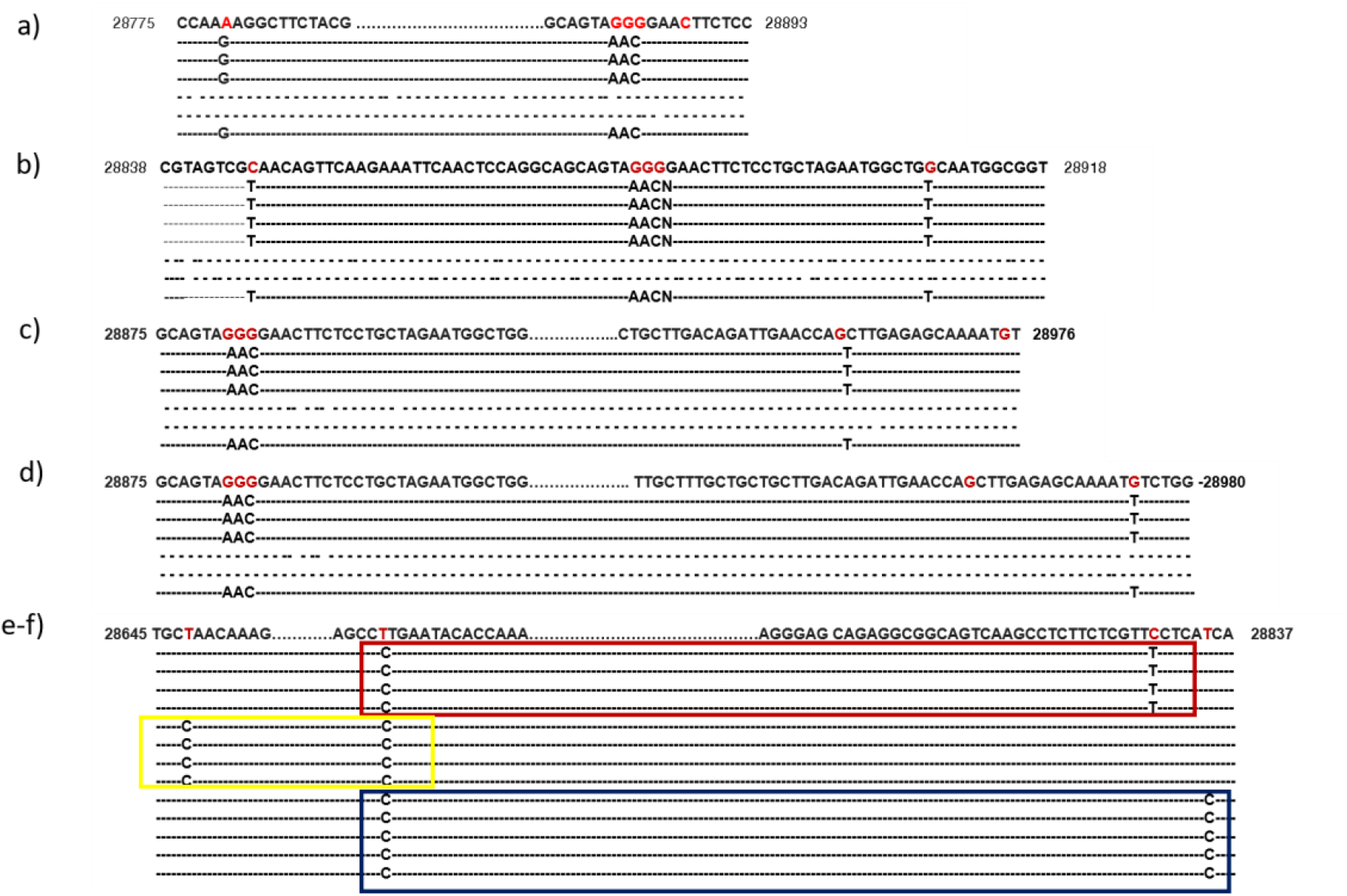
Representation of the haplotype variants was observed in the severe acute respiratory syndrome coronavirus 2 (SARS-CoV-2) genome. a-d) Represent the genomic location of each haplogroup identified in the nucleocapsid region which is a target of RT-PCR based detection of severe acute respiratory syndrome coronavirus 2 (binding sites of primers recommended by the World Health Organization, the Centers for Disease Control and Prevention in USA and China, Germany including Japan’ s Center of Infectious Disease testing protocol. e-g) haplotype variants were observed at nucleocapsid other than the RT-PCR target region.

This protein is associated with the viral structural rearrangement of genomic RNA and serves several functions essential for viral replication and RNA dimerization (13–15). Therefore, these unique haplotype variants may have the possibility to play a role in the variation of pathogenicity, infection rate, recurrence infection, and mortality rate, including the immune response. Therefore, it demands the molecular level study of those haplogroups for their possible association with the parameter mentioned earlier, including mortality rate! Moreover, recently developed vaccines functionality could be validated against those haplogroups from those who did not respond to the given vaccine.

### Major hotspot mutant genes and sites were identified in the global SARS-CoV-2 genome

To identify major hotspot mutants, we analyzed 1,012,582 global SARS-CoV-2 sequence data were available in the NCBI database as of August 4^th^, 2021. We analyzed the global distribution of these sequences for complete and partial genome sequences (Figure 2.) and identified major hotspot mutant genes (Table 2.).

**Table 2.** Major hotspot mutant genes with synonymous mutation, codon, and protein changes with mutation sites of the severe acute respiratory syndrome coronavirus 2 (SARS-CoV-2) genome in 1,012,582 global samples.

| Protein | Protein change | Count (sample) | Genomic location | Codon change | Protein change type |
| --- | --- | --- | --- | --- | --- |
| nsp3 | F106F | 607711 | 3038 | TTT > TTC | synonymous |
| nsp3 | F1089F | 250156 | 5987 | TTT > TTC | synonymous |
| nsp3 | T1189T | 63447 | 6287 | ACT > ACC | synonymous |
| nsp3 | T1607T | 12512 | 7541 | ACC > ACT | synonymous |
| nsp3 | P512P | 10612 | 4256 | CCA > CCG | synonymous |
| surface glycoprotein | I68I | 31555 | 21767 | ATC > ATA | synonymous |
| surface glycoprotein | A924A | 15709 | 24335 | GCT > GCC | synonymous |
| surface glycoprotein | Q613Q | 12596 | 23402 | CAA > CAG | synonymous |
| nucleocapsid phosphoprotein | R203R | 322626 | 28883 | AGA > AGG | synonymous |
| ORF8 protein | H17H | 47834 | 27945 | CAT > CAC | synonymous |
| ORF8 protein | F120F | 10048 | 28254 | TTT > TTC | synonymous |
| nsp6 | V120V | 113439 | 11333 | GTG > GTA | synonymous |
| nsp2 | S36S | 249655 | 914 | TCT > TCC | synonymous |
| nsp2 | N435N | 17370 | 2111 | AAT > AAC | synonymous |
| nsp4 | D144D | 113780 | 8987 | GAT > GAC | synonymous |
| helicase | N268N | 35967 | 17041 | AAC > AAT | synonymous |
| helicase | T137T | 12905 | 16648 | ACT > ACG | synonymous |
| membrane glycoprotein | L93L | 62743 | 26802 | CTG > CTC | synonymous |
| membrane glycoprotein | F53F | 10873 | 26682 | TTT > TTC | synonymous |
| 3'-to-5' exonuclease | D172D | 14808 | 18556 | GAT > GAC | synonymous |
| 2'-O-ribose methyltransferase | A199A | 62438 | 21256 | GCC > GCG | synonymous |

**Figure 2.**
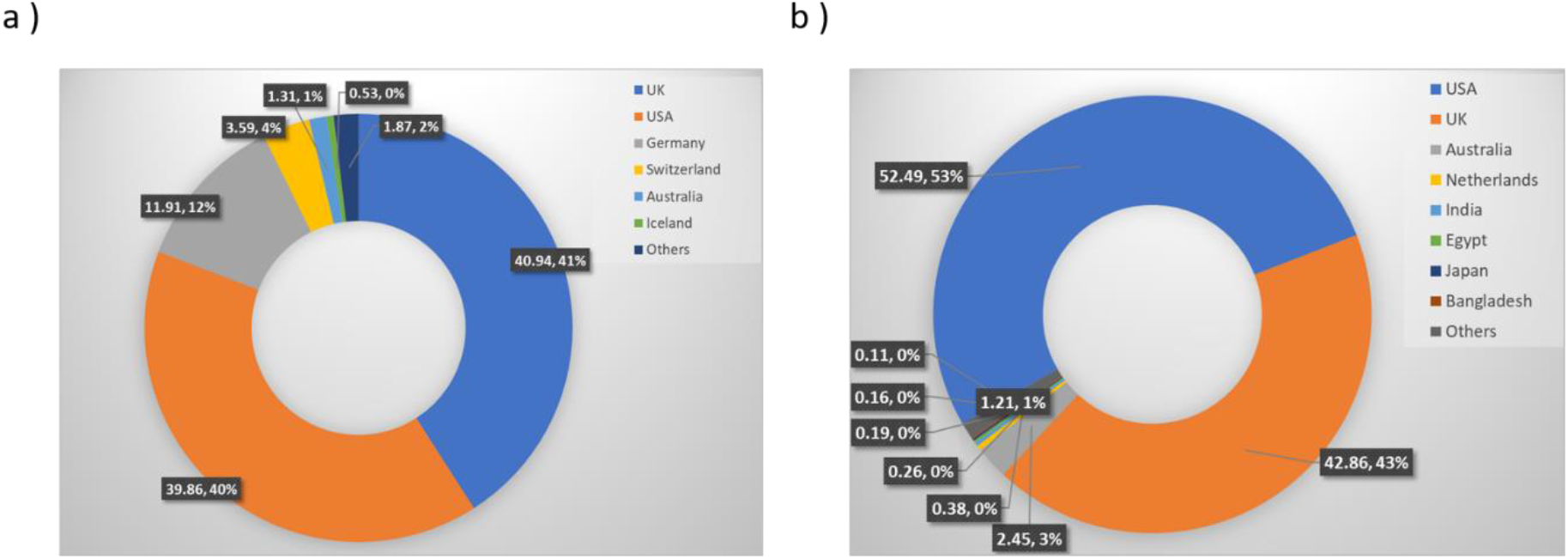
Global distribution of severe acute respiratory syndrome coronavirus 2 (SARS-CoV-2) genomes by sample collection locations from 101 countries. a) Distribution of 1,012,582 SARS-CoV-2 genomes (complete and partial genome sequence) by countries as of August 4^th^, 2021. b) Global distribution of 405,461 complete SARS-CoV-2 genomes by region (from 88 countries).

The global mutation distribution data revealed that the top 11 major mutant genes had synonymous mutations observed in > 50,000 global samples (Table 3.).

**Table 3.** Global distribution of 11 major mutant genes with the number of synonymous and non-synonymous mutations of severe acute respiratory syndrome coronavirus 2 (SARS-CoV-2) genome present at least in >10,000 global samples.

| protein | No. of synonymous Mut. | No. of non-synonymous mut. | Total Mut. |
| --- | --- | --- | --- |
| nsp3 | 5 | 9 | 14 |
| surface glycoprotein | 3 | 22 | 25 |
| nucleocapsid phosphoprotein | 1 | 15 | 16 |
| ORF8 protein | 2 | 7 | 9 |
| nsp6 | 1 | 8 | 9 |
| nsp2 | 2 | 3 | 5 |
| nsp4 | 1 | 4 | 5 |
| helicase | 2 | 2 | 4 |
| membrane glycoprotein | 2 | 1 | 3 |
| 3'-to-5' exonuclease | 1 | 2 | 3 |
| 2'-O-ribose methyltransferase | 1 | 1 | 2 |

Mutation at each of the surface glycoprotein, nucleocapsid phosphoprotein, ORF8, and ORF3a protein-coding gene was observed in 24%, 19%, 7%, and 3% of the global samples respectively, while top mutations at non-structural protein-coding genes nsp3 and at each of the nsp6, nsp4 and nsp2 were observed in 18% and 4% of the global samples respectively. In addition, ORF3a protein and Helicase coding genes were observed to have a mutation at 3%, while both ORF7a and 3’-5’ exonuclease were observed to have a mutation in 2% of the global samples (Figure. 3).

**Figure 3.**
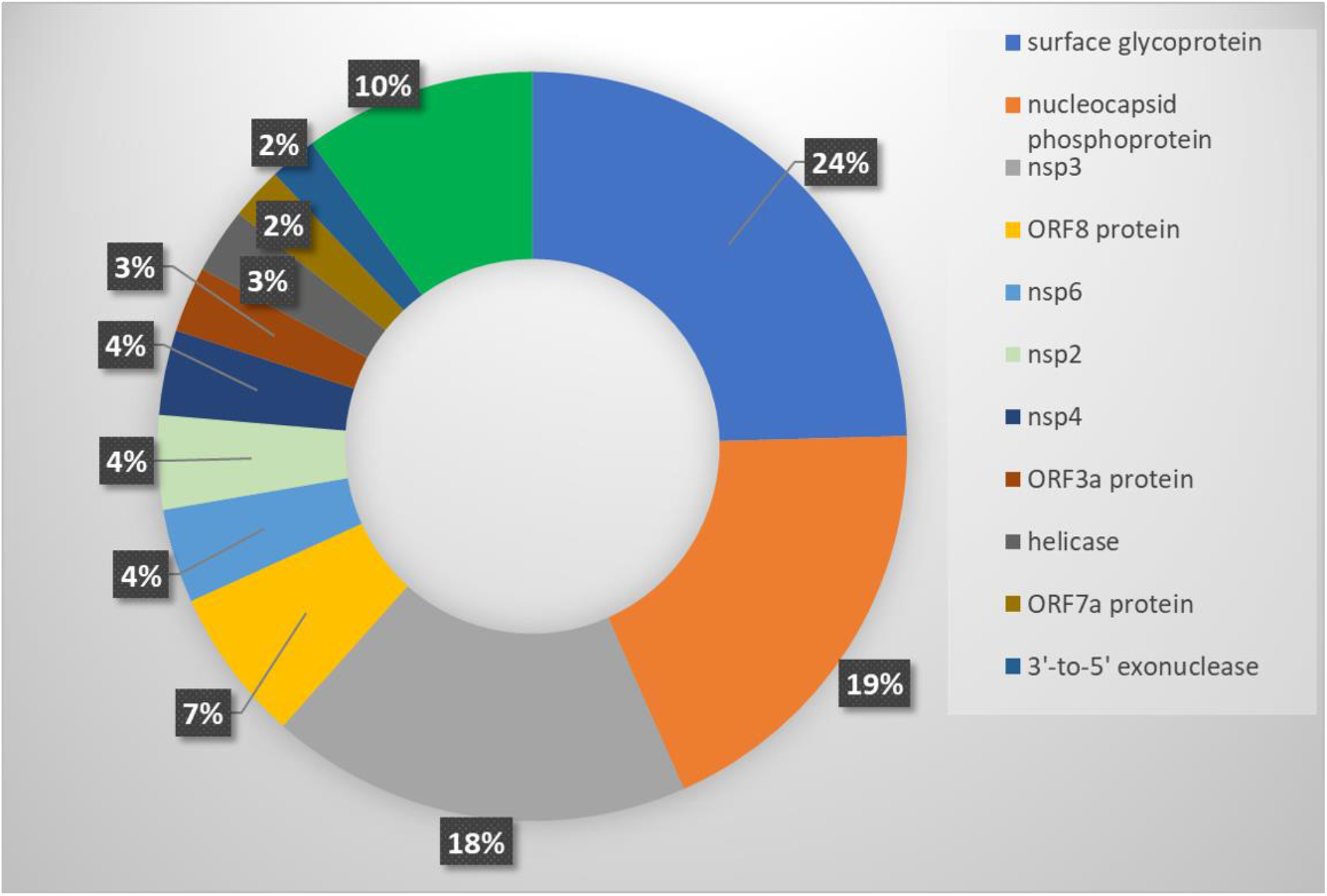
Distribution of major hotspot mutant genes in 405,461 global samples with complete SARS-CoV-2 genome sequences. Complete genomes sequences from National Center for Biotechnology Information (NCBI) database were analyzed for top mutant genes as of August 4^th,^ 2021, a total of 405,461 complete SARS-CoV-2 genomes from 88 countries were analyzed.

## Discussion

In this multi-population-based SARS-CoV-2 genome analysis study, we have identified six unique haplotype variants and 11 major hotspot mutant genes that might play a crucial role in inter-individual variation in COVID-19 pathogenicity, severity, immune response, and mortality rate. Studying the association of these variants and hotspot mutant genes at the cellular and molecular level may help understanding the mechanisms of pathogenicity, progression of the disease to more severity and mortality, response to drugs, and immune response to vaccines. Therefore, it will help manage individual SARS-CoV-2 patient groups with identified haplotype variants and major mutant genes by developing effective drugs and vaccines for the target subgroups. In addition, we have identified SARS-CoV-2 clones with mutations at primer and probe binding sites that might cause false-negative PCR detection results. Efficient diagnosis and treatment strategies have become a big challenge to the medical community and healthcare professionals due to the significantly high false-negative detections rate. We showed in our previous study that high false-negative results might be due to the genomic variations at the primer and probe binding sites. In the current study, we have identified global SARS-CoV-2 clones with mutations at target PCR-detections sites. The mutations were observed at either primer-probe binding sites or both sites (Table 1.). We also observed that some clones showed mutations at the multiple primer-prob sites recommended by CDC, USA, China and Japan’s NIID (Supplementary Table 1.) raises the concern. These concerns have emerged recently, notably regarding sensitivity and accuracy of the RT-PCR-based detection of false-negative data even after frequent retesting procedures, and might play a significant role in transmitting the virus without traceability of the sources. Therefore, we recommend using at least three more alternative primer-probe sets for RT-PCR detections of SARS-CoV-2 along with the currently used primers and probes sets.

While we were analyzing the variants at the target detections sites, we identified six unique haplotype variants, at the nucleocapsid regions, N encoding nucleocapsid phosphoprotein of which three variants are present at or near the target detection sites; however, the other three haplotype variants are located at the distant upstream of the target detection sites (Figure 2.). Nucleocapsid phosphoprotein (N), also known as the replication-transcription complexes (RTCs), has been reported to be associated with early and late viral replication, structural rearrangement of the genomic RNA, viral RNA dimerization and serves several functions essential for viral replication (31–34). Therefore, it demands the molecular level studies if these haplotype variants present in target subgroups are associated with the alterations of nucleocapsid functions in SRAS-CoV-2 pathogenicity and if they facilitate the functions of other structural or non-structural proteins. We also investigated other genes that have been reported to exhibit functions in viral pathogenesis and are the targets for anti-viral drug development (35–43). We identified eleven major mutant genes with major hotspot and synonymous mutations, each of which mutations were observed in at least 50,000 global samples (Table. 2). The global distribution of these major mutant genes revealed that the highest mutations were present in structural protein-coding gene surface glycoproteins, nucleocapsid phosphoprotein, and non-structural coding gene nsp3 with 25, 16, and 9 hotspots mutant sites, respectively, in the global samples. Mutations identified in surface glycoprotein could affect its function in the receptor recognition and cell membrane fusion process with host-receptors angiotensin converting enzyme 2 (ACE2) (42–44).

Synonymous and non-synonymous mutation detected in the nucleocapsid phosphoprotein region resides at SARS-CoV-2 RNA synthesis sites might have a negative regulatory influence in viral genomic RNA packaging during virion assembly and suppression of host immune response through RNA-dependent phase separation (45–46). In addition, the C-terminal domain of nucleocapsid phosphoprotein has been reported to be associated with anchoring the viral Nsp3, also known as papain-like protease, a component of RTCs. nsp3 catalyzes the reaction that preferentially cleaves ubiquitin-like interferon-stimulated gene 15 (ISG15) protein from interferon factor 3 (IRF3) which weakens the type I interferon response, could exacerbate hyperinflammatory conditions and progression to severe COVID-19 (47–48). nsp2 and nsp3 are conserved sequences that have no homology with other Coronaviruses. Moreover, ORF8, and 3’- to- 5’ exonuclease (nsp14) has been reported to suppress immune response through disrupting IFN-I signaling, down-regulating MHC-I, and inhibiting IFNγ-induced anti-viral gene expression in human lung epithelial cells (49–52) while membrane glycoprotein (M) has been reported to acts as a negative regulator of innate immune response (31–32, 53).

Therefore, mutation analysis of these genes may reveal potential mechanisms that distinguish COVID-19 from other viruses, as well as inter-individual differences in immune response and COVID-19 severity.

Helicase (nsp13), the most conserving site of SARS-CoV-2, contains two druggable pockets, nucleoside triphosphate hydrolase (NTPase) and helicase activities that hydrolyze and unwind RNA helices. In viral life cycles, nsp13 and nsp14 play the central role in RNA replication by unwinding the duplex RNA and its exoribonuclease (ExoN) N7-methyltransferase (N7-MTase) activities, respectively. In addition, Nsp13 facilitates the correct folding of the viral protein into 2ndary and tertiary structures to become functional. Therefore, studying the mutations in this gene could suggest possible interindividual variation in the drug response and pathogenicity (52, 54–56).

To understand the COVID-19 related target drug-gene interactions and for the selection of effective drugs, molecular level studies will be needed for each of the proposed target variants. Any target drug or chemical compound should be molecularly docked for its binding affinity with the proteins of the host cells for example angiotensin converting enzyme II (ACE II) as well as with the proteins expressed by the target genes of SARS-CoV-2 genome. In addition, studying the molecular network or signaling will be needed. Furthermore, investigating if these unique variants will have impacts on drug-gene interaction, signaling network, as well as impacts on pharmacokinetics using target chemical compounds that are used to treat COVID-19 patients, for example, Diosgenin, Syringaresinol-O-beta-D-glucoside, etc., are present in the traditional Chinese medicinal herb used to treat COVID-19 patients as an alternative could be a subject for future studies (57, 58).We did analyze the number of mutations and sites of mutations of each of the crucial eleven mutant genes (Table 3). To avoid the biases of the sequence data, sequencing procedures including PCR-based sequencing and machines and analysis pipeline may cause errors we avoided genes that have been found mutated in < 10,000 global samples (supplementary table 2). For the first time, we are reporting the unique haplotype variants and other potential targets variants in 11 major mutant genes by analyzing a large number of SARS-CoV-2 global samples (n=1, 012,582). A comparison of the analytics has been performed in the present study with the one existing similar investigation (Table 4.) demonstrating the importance of the present study.

**Table 4.** Comparison of analytics of the present study with the existing similar literature. A comparison of analytics was made representing the global sample volume, distribution, unique variants, major mutant genes, variant sites, and filtering criteria.

| Types of analytics | The one existing similar literature | In the current study |
| --- | --- | --- |
| Number of genomes analyzed | 3067 | 1, 012,582 |
| Global distribution of host | 55 countries | 88 countries |
| Unique haplotype variants | No such studies | 6 unique variants |
| Number of hotspot mutant genes | 6 | 11 |
| Total variant sites | 782, of which 582 (65 %) had non-synonymous effects | 21, 509, of which 12,910 (60%) had non-synonymous effects |
| Technical error filtering (exclusion of samples having a mutation in 10,000 samples, < 2%) for major mutant genes | No | Yes |
| Information on mutation sites on major mutant genes (synonymous or non-synonymous) | No | yes |

All these crucial mutant genes have been reported to be linked to SARS-CoV-2 pathogenicity, viral replication, virus-host interaction, transmission, and immune response to the host (42–56). Therefore, any individual subgroups with these mutations may have shown variations in gene functions and mechanisms mediating the traits or phenotypes caused by mutations and may require special management procedures, treatment strategies, and effective vaccinations. Further molecular level studies are needed to investigate the effects of these mutations.

## Conclusion

Genome analysis data of our study may play a significant role in understating interindividual variations in drug response and immune response by vaccines and variations in the pathogenicity, recurrence of infection, and mortality among nations and subgroups.

## Data Availability

All data generated or analyzed during this study are included in this published article (supplementary information files).

---

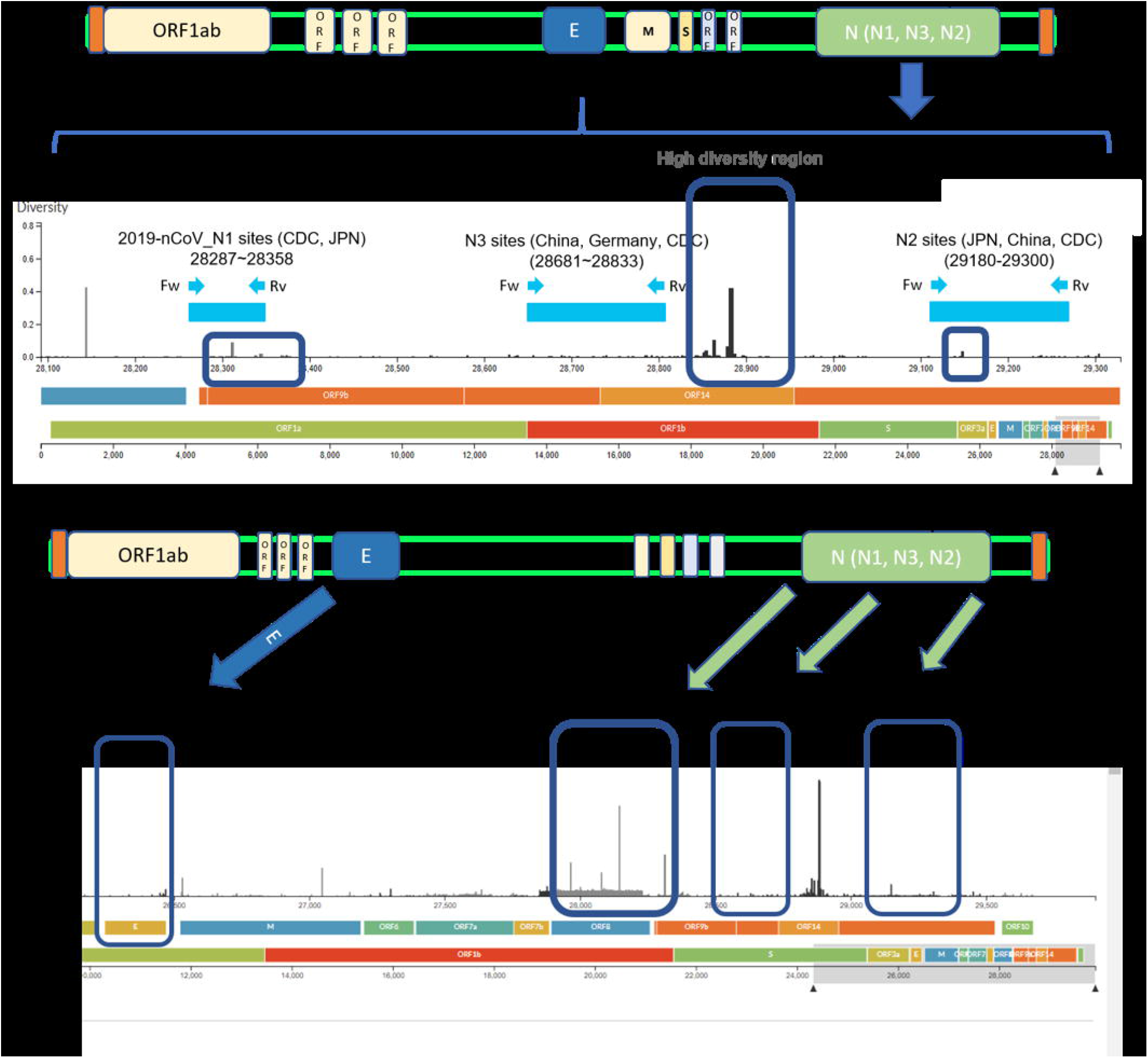

